# Systematic assessment of early brain injury severity at admission with aneurysmal subarachnoid hemorrhage

**DOI:** 10.1101/2023.10.17.23297185

**Authors:** Sheri Tuzi, Beate Kranawetter, Dorothee Mielke, Veit Rohde, Vesna Malinova

**Author notes:** **Corresponding author: Vesna Malinova**, MD Department of Neurosurgery Georg-August-University Robert-Koch-Straße 40 37075 Göttingen.

## Abstract

**Background:** Early brain injury (EBI) after aneurysmal subarachnoid hemorrhage (aSAH) has been increasingly recognized as a risk factor for delayed cerebral ischemia (DCI). While several clinical and radiological EBI biomarkers have been identified, no tool for systematic assessment of EBI severity has been established so far. This study aimed to develop an EBI grading system based on clinical signs and neuroimaging for estimation of EBI severity at admission.

**Methods:** This is a retrospective observational study assessing imaging parameters (intracranial blood amount, global cerebral edema (GCE)), and clinical signs (persistent loss of consciousness [LOC]) representative for EBI. The intracranial blood amount was semi-quantitatively assessed. One point was added for GCE and LOC, respectively. All points were summed up resulting in an EBI grading ranging from 1-5. The estimated EBI severity was correlated with progressive GCE requiring decompressive hemicraniectomy (DHC), DCI-associated infarction, and outcome according to the modified Rankin scale (mRS) at 3-month-follow up.

**Results:** A consecutive cohort including 324 aSAH-patients with a mean age of 55.9 years, was analyzed. The probability of developing progressive GCE was 9% for EBI grade 1, 28% for EBI grade 2, 43% for EBI grade 3, 61% for EBI grade 4, and 89% for EBI grade 5. The EBI grading correlated significantly with the need for DHC (r=0.25, *p*<0.0001), delayed infarction (r=0.30, *p*<0.0001), and outcome (r=0.31, *p*<0.0001).

**Conclusions:** An EBI grading based on clinical and imaging parameters allowed an early systematic estimation of EBI severity with a higher EBI grade associated not only with a progressive GCE but also with DCI and poor outcome.

## Introduction

Aneurysmal subarachnoid hemorrhage (aSAH) is a serious medical condition with high morbidity and mortality rates despite continuous improvements in neurocritical care over the years. Early brain injury (EBI) has been increasingly recognized for its substantial contribution to poor outcome after aSAH. Furthermore, EBI has been also shown to be a risk factor for delayed cerebral ischemia (DCI)^1^. The term EBI summarizes multifactorial pathomechanisms starting with aneurysm rupture and manifesting within the first 72 hours after ictus^2^. Neuronal cell death leads to blood-brain-barrier breakdown resulting into global cerebral edema (GCE) and microcirculatory dysfunction, which belong to the most frequent consequences of EBI^3^. Although there is growing evidence about EBI from experimental studies, these concepts have not been implemented in clinical practice yet. Several grading scales are routinely used to capture the severity of aSAH focusing on clinical (WFNS = World Federation of Neurosurgical Societies scale) or radiological (Fisher scale) parameters. Although these scores are representing the severity of EBI to some degree, they were not specifically created for the assessment of EBI. Recently, a score based on early edema signs on initial computed tomography scan (SEBES = Subarachnoid hemorrhage early brain edema score) was published for evaluation of EBI severity^4^. Ictal loss of consciousness (LOC) and total intracranial blood burden are another frequently used surrogate markers for EBI^5^. A reliable EBI grading is needed to allow a better risk stratification for secondary complications of severe EBI like progressive GCE requiring decompressive hemicraniectomy (DHC). Nevertheless, no such tool for systematic EBI assessment including clinical and radiological surrogate parameters for EBI has been established yet. The aim of this study was to develop an EBI grading based on clinical and radiological parameters on admission and to evaluate its predictive value for progressive GCE, delayed infarction, and functional outcome after aSAH.

## Materials and methods

### Patient population

This is a retrospective observational study. A consecutive patient cohort diagnosed with aSAH and treated at our Department of Neurosurgery during the period from January 2012 to December 2020 was analyzed. Diagnosis of aSAH was confirmed by cranial computed tomography (CCT), computed tomography angiography (CTA), and/or digital subtraction angiography (DSA), which confirmed the presence of a ruptured aneurysm. Patients presenting with non-aneurysmal SAH were excluded from the study. The study was performed in accordance with our institution’s ethical committee and the Helsinki declaration. The study was approved by the local institutional review board the Ethics Committee of the University Medical Center Göttingen (Number 16/9/20). The manuscript was written according to the STROBE guidelines for reporting observational cohort studies.

### EBI grading calculation

The severity of EBI was assessed based on the following parameters: persistent loss of consciousness (LOC) upon admission (patients who were only unconscious for a short time due to an epileptic seizure were not considered to have persistent LOC), GCE defined as SEBES ≥ 3, and the total intracranial blood burden. One point was assigned for persistent LOC, and one point for GCE on admission CT. The intracranial blood burden was semi-quantitatively assessed using three scoring systems reflecting the blood amount within different intracranial areas (subarachnoid space, ventricular system, and intraparenchymal hematoma). The Hijdra score^6^ was used to assess the subarachnoid blood amount, the Le Roux score^7^ was applied to capture the intraventricular blood amount, and the intraparenchymal blood volume was assessed using the ABC/2 ellipsoid formula^8^. The Hijdra score stratifies the amount of extravasated blood in each of 10 basal cisterns and fissures, and ranges from 0 (no blood) to 3 (full of blood), with a maximum score of 30 points. The Le Roux score reflects the amount of blood in each of the four ventricles, and ranges from 0 (no blood) to 4 (filled with blood and expanded), with a maximum score of 16 points. The intraparenchymal blood volume was assessed using the ABC/2 formula, and a point system was assigned based on the hematoma volume: 1 point for <10 ml, 2 points for 10-30 ml, and 3 points for >30 ml. The maximal achievable total intracranial blood burden score was 49 points, which was calculated by summing up the scores assigned for the subarachnoid blood amount, intraventricular blood amount, and intraparenchymal blood amount, respectively. The total intracranial blood burden was divided into three categories: low (one point was assigned for low intracranial blood burden), moderate (two points were assigned for moderate intracranial blood burden), and high (three points were assigned for high intracranial blood burden). Low intracranial blood burden corresponded to scores between 0 and 19 points, moderate intracranial blood burden was assigned for scores of 20-29 points, and high intracranial blood burden was considered whenever scores of 30 points or more were reached. The sum of all assigned points resulted in the EBI grade raging from 1 to 5. To consider the evolution of EBI in the first three days after ictus, the EBI grade was calculated on admission and on day 3 after aneurysm rupture.

### Primary and secondary outcome parameters

The primary outcome parameter was the need for DHC due to progressive GCE with a refractory ICP increase, which is regarded as a severe manifestation of EBI. The GCE was assessed based on the CT scans on day 1 and day 3 applying the SEBES score^4^, where SEBES score of at least 3 was regarded as GCE. ICP was monitored via an intraparenchymal probe (usually placed within the right frontal lobe) in comatose or sedated patients. Secondary outcome parameters were delayed infarctions and the functional outcome according to the modified Rankin scale (mRS) at the 3-month follow-up examination by the neurosurgeon in the outpatient care unit. The mRS ranges from 0 (no symptoms) to 6 (death). Delayed infarctions were defined as newly diagnosed cerebral infarctions after excluding treatment-associated infarctions by performing a CT scan within 24 hours after the procedure of aneurysm treatment (clipping or coiling).

### Statistical analysis

The statistical analysis was conducted using GraphPad Prism software (Version 9, GraphPad Software, San Diego, CA, USA). Descriptive and inferential statistics were utilized to describe and analyze the collected data. Frequency and percentages were used to depict discrete, ordinal, and binary variables, while mean ± standard deviation was used to depict continuous variables. For each parameter, a confidence interval (CI) was determined. Spearman’s rho correlation coefficient was applied to assess the correlation between two variables. The sensitivity and specificity, as well as the negative and positive predictive values, were calculated using a 2 x 2 contingency table. The significance, or p-value, between the two variables was then evaluated using the Fisher exact test. Receiver operating characteristic curve (ROC curve) analysis was performed to numerically display the relationship between sensitivity and specificity, and to estimate the overall diagnostic accuracy of the tests.

## Results

### Patient characteristics

A consecutive patient cohort of 324 patients with aSAH was included in this study. The cohort consisted of 206 (64%) female and 117 (36%) male patients, with a mean age of 55.9 ± 13.5 years, ranging from 23 to 90 years. The ruptured aneurysm was located within the anterior circulation in 277 (86%) cases, and 46 (14%) cases had a ruptured aneurysm of the posterior circulation in. A poor WFNS grade (IV-V) had 44% (143/324) of the patients. The aneurysm was treated by microsurgical clipping in 172 (53%) cases and by endovascular coiling in 152 (47%). An overview of the patient characteristics is presented in **Table 1**.

**Table 1.** Baseline Characteristics.

|  |  |
| --- | --- |
| **Number of patients** | 324 |
| **Mean age (in years)** | 55.9 SD 13.5 |
| **Sex** |  |
| - Male | 117 (36%) |
| - Female | 206 (64%) |
| **Aneurysm location** |  |
| - Anterior circulation | 277 (86%) |
| - Posterior circulation | 46 (14%) |
| **Aneurysm treatment** |  |
| -clipping | 172 (53%) |
| -coiling | 152 (47%) |
| **WFNS I-III** | 182 (56%) |
| **WFNS IV-V** | 142 (44%) |
| **Fisher score 1-2** | 29 (9%) |
| **Fisher score 3-4** | 295 (91%) |

### EBI grading vs. progressive GCE requiring decompressive hemicraniectomy

The distribution of EBI grades on day 1 and day 3 is summarized in **Table 2**. Severe EBI (EBI grade 4 and 5) was found in 19% of patients on day 1 and in 21% of patients on day 3. Progressive GCE with need for DHC occurred in 13% (41/324) of all patients. Regarding the EBI grading, progressive GCE requiring DHC was found in 89% of patients with EBI grade 5, in 61% of patients with EBI grade 4, in 43% of patients with EBI grade 3, in 28% of patients with EBI grade 2, and in 9% of patients with EBI grade 1. An EBI grade of ≥3 on day 3 correlated significantly with the need for a DHC (AUC 0.8, *p*<0.0001 (**Figure 1**). The negative predictive value was 98% (95%CI 95-99%) (**Figure 1**). The EBI grading on day 3 showed a high sensitivity of 93% (95%CI 81-97%) and a high odds ratio of 17.2 (95%CI 5.7-53.9) for identifying patients, who are at risk for develop progressive GCE requiring DHC (**Figure 1**).

**Table 2.** Distribution of EBI grading on day 1 and on day 3 after ictus.

| EBI grading | 24 hours N (%) | 72 hours N (%) |
| --- | --- | --- |
| EBI grade 1 | 55/324 (17%) | 54/324 (17%) |
| EBI grade 2 | 138/324 (42%) | 111/324 (34%) |
| EBI grade 3 | 71/324 (22%) | 92/324 (28%) |
| EBI grade 4 | 47/324 (15%) | 46/324 (14%) |
| EBI grade 5 | 13/324 (4%) | 21/324 (7%) |

**Figure 1.**
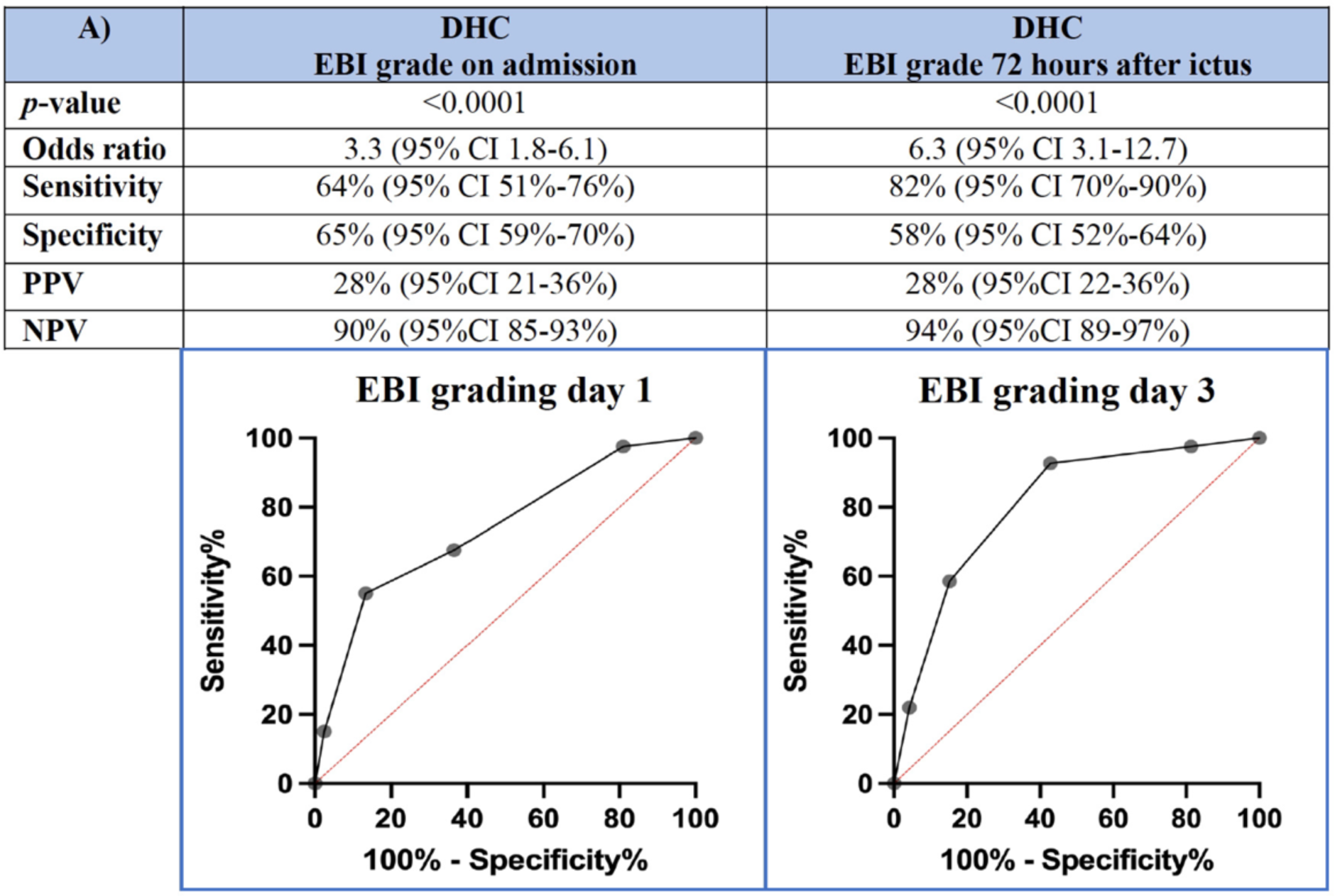
Discrimination power (ROC analysis) of EBI grading regarding progressive global cerebral edema requiring decompressive hemicraniectomy on admission (day 1) and 72 hours after ictus (day 3), respectively.

### EBI grading vs. delayed infarctions and functional outcome

Delayed infarctions occurred in 17% (56/324) of all patients. Severe EBI (EBI grade ≥3) was associated with a higher incidence of delayed infarctions than patients with an EBI grade <3, which was the case on day 1 (AUC 0.72 (**Figure 2**) as well as on day 3 (AUC 0.75 (**Figure 2**). An EBI grade of ≥3 correlated significantly with delayed infarctions both on day 1 (p<0.0001) and day 3 (<0.0001). The negative predictive value was 90% (95%CI 85-93%) on day 1 and 94 % (95%CI 89-97%) on day 3 (**Figure 2**).

**Figure 2.**
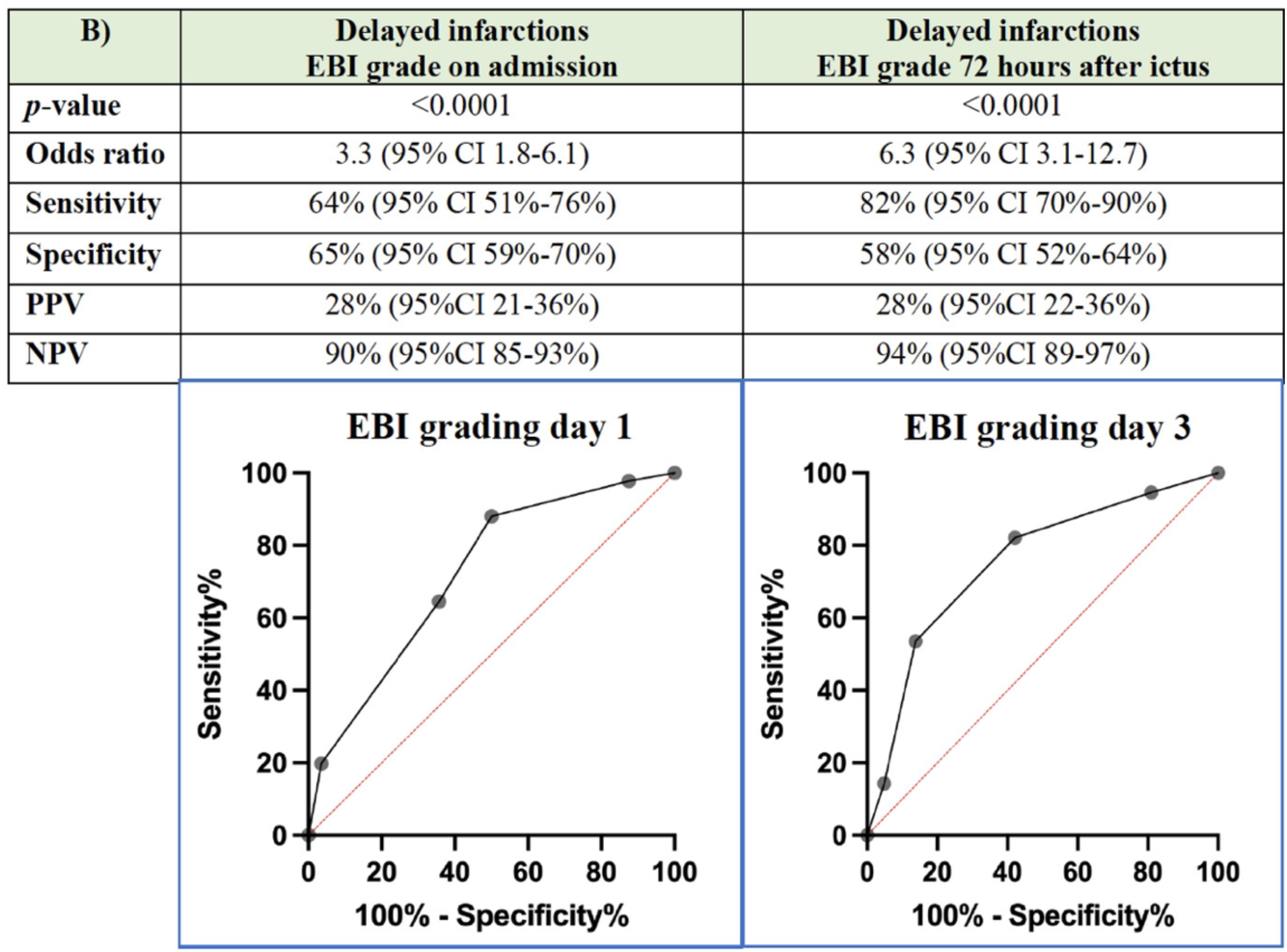
Discrimination power (ROC analysis) of EBI grading regarding the development of delayed infarctions on admission (day 1) and 72 hours after ictus (day 3), respectively.

The average mRS in the patient cohort was 1.6 (SD 1.9) at 3-months follow-up. Patients with an EBI grade ≥ 3 had experienced significantly more often poor funtional outcome (mRS >3) both on day 1 (p<0.0001, AUC 0.67) and day 3 (p<0.0001, AUC 0.70 (**Figure 3**). The negative predictive value was 83% (95%CI 76-87 %) on day 1 and 85% (95%CI 79-90%) on day 3 (**Figure 3**).

**Figure 3.**
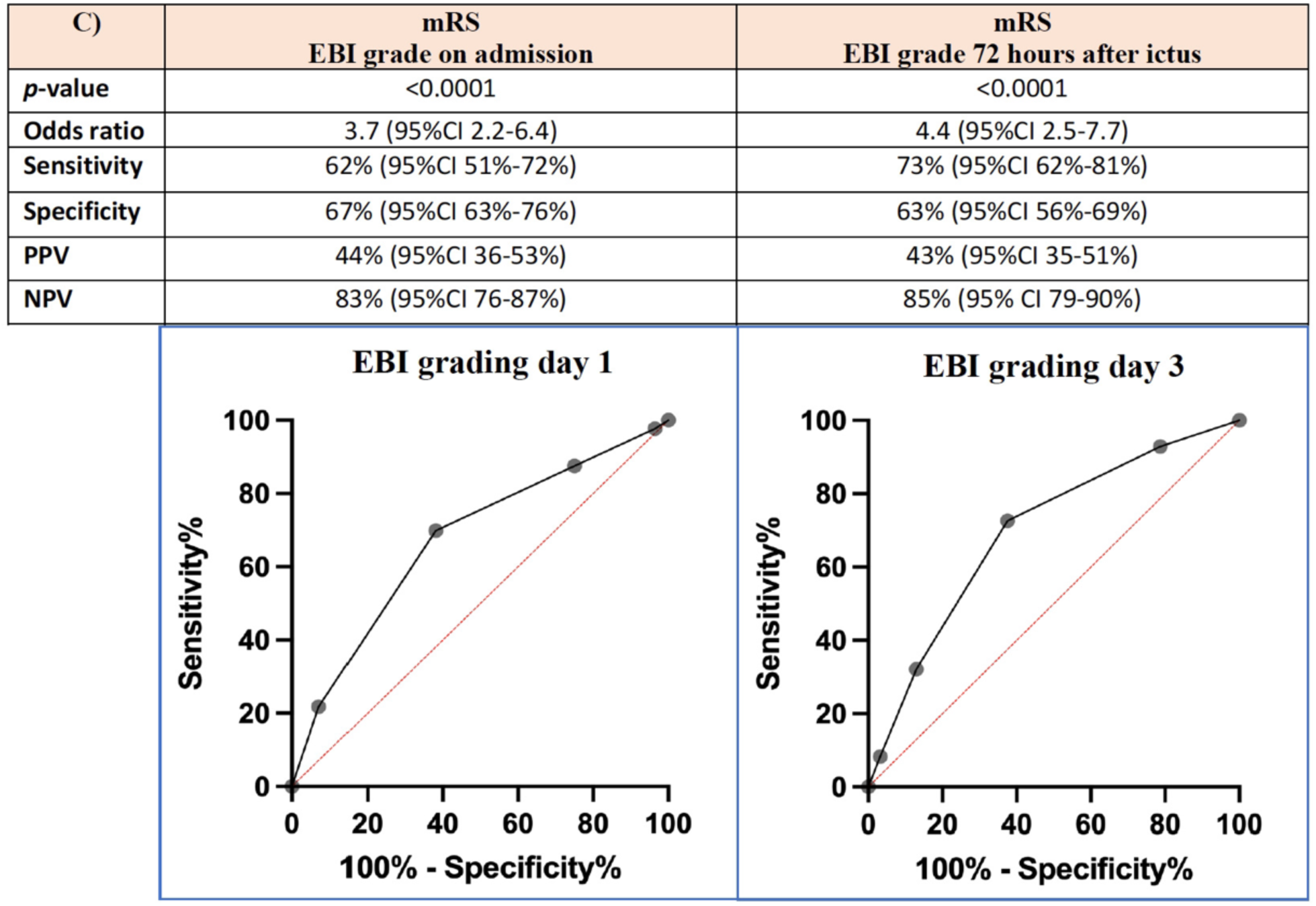
Discrimination power (ROC analysis) of EBI grading regarding the functional outcome according to modified Rankin scale (mRS) at 3-months follow up on admission (day 1) and 72 hours after ictus (day 3), respectively.

## Discussion

An intracranial aneurysm rupture initiates multiple pathophysiological processes happening within the first 72 hours after ictus, which are summarized with the term EBI. These processes have been demonstrated not only to correlate with early and delayed complications of the bleeding, but also to have an impact on functional outcome after aSAH. Despite growing evidence supporting the prognostic value of EBI no grading system has been established so far to systematically assess EBI severity in clinical practice. This study strived to fill this gap and to develop a grading system that stratifies the severity of EBI considering clinical and radiological parameters, which serve as surrogate markers for EBI: persistent LOC as a clinical sign of substantial brain injury, presence of GCE corresponding to SEBES ý3^4^, and the total intracranial blood burden considering different intracranial compartments like subarachnoid space, ventricular system, and brain parenchyma^6–8^. All three EBI surrogate markers are available on admission after aneurysm rupture allowing an EBI grading early on following aSAH diagnosis. Since EBI is a dynamic process, EBI grading was calculated at two time points (wihtin 24 and 72 hours) after aSAH-diagnosis to consider the dynamics of these parameters within the first 72 hours after ictus. Both assessment time points are providing different information concerning the pathophysiological processes happening within the first days after aSAH. While the EBI grade on admission is deemed to allow an early prediction of expected EBI severity bearing higher risk for secondary complications, the EBI grade 72 hours after ictus is more a depiction of the already manifested EBI severity.

### Surrogate markers of EBI

The intracranial blood burden is an important indicator of aSAH severity. The amount of extravasated blood in the intraparenchymal, intraventricular, and subarachnoid space, plays an important role in predicting DCI after aSAH^2^. The Hijdra score^6^ stratifying the amount of subarachnoid blood has been shown to correlate with the risk of developing cerebral vasospasm^9^. On the other hand, the LeRoux score evaluates the intraventricular blood amount correlated with the functional outcome of patients with interventricular hemorrhage^10,11^. The total intracranial blood burden plays an important role in the development and progression of EBI due to the neurotoxicity of hematoma components^12^. Hemoglobin released during hematoma degradation leads to inflammation, oxidation, nitric oxide release, and edema, aggravating neuronal injury^13^. Higher and earlier blood clearance has been associated with better overall outcome^14^. Since hematoma components have been directly linked to EBI, reducing the intracranial blood burden, and promoting a fast blood clearance could potentially ameliorate the severity of EBI^12^. An experimental study suggested a strong correlation between subarachnoid blood clots and GCE^15^, further reinforcing the idea that higher blood clearance and lower intracranial blood burden could mitigate the progression of EBI potentially resulting into progressive GCE, which in turn is correlated to poor outcome^16^.

### Diagnostic and therapeutic implications of using an EBI grading in clinical practice

Progressive GCE resulting into increased ICP, that if refractory to conservative treatment requires neurosurgical intervention like DHC represents one of most severe EBI manifestations after aSAH. The EBI grading presented here showed a significant correlation with the development of progressive GCE requiring DHC. According to these findings, an EBI grading calculated on admission seems to facilitate a reliable and early stratification of EBI severity, separating patients with high risk for progressive edema needing DHC from those with low risk of developing this severe complication. Currently, the indication for DHC in aSAH patients is set on an individual basis with DHC usually performed in case of increased ICP refractory to maximal conservative treatment. The role of early primary DHC has not been defined yet and is currently under investigation in a prospective randomized controlled trial (PICASSO)^17^.

An early primary DHC seems to improve the functional outcome and reduce the mortality rate in poor-grade aSAH^18^. However, there is no clear evidence that patients with cerebral edema or infarction might benefit from DHC. Two recently published studies support the concept that an early DHC might be more protective than a later one, as it might potentially reduce ICP and not confine the post-hemorrhagic swelling, thus leading to better oxygenation and overall better brain tissue perfusion^19,20^. Even in an equally distributed clinical status, i.e., disregarding good-and poor-grade aSAH, patients showed more favorable outcomes and seemed to have a long-term benefit from DHC. Even a prophylactic DHC could potentially be beneficial^19^. Jabbarli et al. have proposed a risk score for DHC after aSAH (PRESSURE score) including several parameters such as high Hunt & Hess grade, younger age (< 55 years), early vasospasm, presence of intracerebral hemorrhage, aneurysm > 5 mm, aneurysm clipping, and the presence of a ventricular drain. A higher score was associated with a higher risk for DHC and lower mortality and morbidity for the patient group with ultra-early DHC performed within 24 hours after ictus^21^. However, none of the previously published studies have considered an EBI-specific grading system in this context. So far, no treatment algorithm exists for the treatment of aSAH according to EBI severity. An EBI grading system would facilitate the consideration of EBI during the decision-making concerning the timing and initiation of further diagnostic and therapeutic measures, which may optimize the clinical management of aSAH patients by allowing a more peronalized treatment planning. The EBI grading significantly correlated not only with the indication for DHC, but also with the development of DCI, and with functional outcome at the 3-months follow-up. Patients presenting with EBI grading of ≥ 3 need to be monitored more carefully and individual clinical management is important for the prevention of severe complications that would lead to worse functional outcome. However, this study does not have the power and was not conducted to answer the question of DHC-timing after aSAH. The establishment of an EBI severity grading may also be helpful for a more reliable patient selection as an inclusion criterion in studies evaluating therapies for EBI not only surgical but also medical treatments such as anti-inflammatory agents, which is the objective of the currently ongoing FINISHER trial^22^.

### Limitations of the study

Limitations of this study include the fact that this is a retrospective study, and the data collected has not been specifically designed for it. Also, the calculation of the intracranial blood burden, while simple, is slightly time-consuming, taking about two minutes to perform, and subjective to the observer. While there is a clear correlation between the EBI grading and the need for DHC, the development of DCI and the overall functional outcome, this grading-system needs to be validated in a multicentric, prospective study to be established as a proper EBI severity assessment tool.

## Conclusion

This is the first study so far which stratifies and assesses the EBI severity in a clinical setting, therefore paving the way for a more targeted clinical management of each individual patient case. A systematic evaluation of LOC, GCE, and intracranial blood burden allowed a valid stratification of EBI severity on admission in aSAH patients. Higher EBI grades were associated with progressive GCE requiring DHC in the early phase after aSAH, higher incidence of delayed infarctions, and poor functional outcome.

## Data Availability

All data available without compromising individual privacy is already presented in the manuscript.

## Acknowledgements

None.

